# Sensorimotor effects of heatwrap and exercise in acute low back pain: results of a randomised controlled trial

**DOI:** 10.64898/2026.08.31.26361841

**Authors:** Claudia Côté-Picard, Amélie Desgagné, Jean Tittley, Catherine Mailloux, Kadija Perreault, Catherine Mercier, Clermont E. Dionne, Jean-Sébastien Roy, Hugo Massé-Alarie

**Affiliations:** Centre interdisciplinaire de recherche en réadaptation et intégration sociale (Cirris), Centre intégré universitaire de santé et de services sociaux (CIUSSS) de la Capitale-Nationale, Québec, Canada; École des sciences de la réadaptation, Université Laval, Québec, Canada; Centre de Recherche du CHU de Québec - Université Laval, Québec, Canada

## Abstract

**Background:** Heatwrap is recommended for acute low back pain (ALBP), and previous research found heatwrap plus exercise more effective than each intervention alone. While recommended by clinical guidelines, their impact on mechanistic outcomes is unknown. This trial aimed to (i) assess immediate and short-term effects of heatwrap alone or combined with exercise, compared with a sham heatwrap, on pain sensitivity, lumbar muscle activity, current pain intensity, and trunk flexion range of motion, and (ii) explore whether changes in pain sensitivity and lumbar muscle activity are associated with changes in clinical symptoms from baseline to 1-week follow-up.

**Methods:** A randomised controlled trial took place at a single research center. Of 315 individuals screened for eligibility, 99 adults with ALBP were recruited and assigned to one of three intervention groups: heatwrap plus exercise (n=34), heatwrap alone (n=33) or sham heatwrap (n=32). Interventions were applied for one hour at the first visit, and immediate effects were measured. Then, interventions were applied for 7 days, and short-term effects were measured at 1-week follow-up. Outcomes included pressure pain threshold, temporal summation of pain, flexion-relaxation ratio, trunk range of motion and current pain intensity.

**Results:** Heatwrap and exercise did not produce greater effects over time than heatwrap alone or a sham heatwrap on all outcomes, and changes in sensorimotor outcomes at one week were not associated with changes in symptoms.

**Conclusions:** Heatwrap and/or exercises did not influence specifically the potential sensorimotor mechanisms tested in individuals with ALBP.

**Trial registration:** ClinicalTrials.gov; registration number: NCT03986047

## Introduction

Low back pain (LBP) is the leading cause of disability globally, affecting more than 619 million people.^1^ The natural evolution of acute LBP (ALBP) is usually favourable in the first 6 weeks, but residual pain and disability remain after a year.^2^ Heatwrap is recommended as first-line care in ALBP, based on evidence that it provides short-term pain relief.^3–5^ A 2005 randomised controlled trial (RCT) found that combining heatwrap and exercise improved acute and subacute LBP more than either intervention alone or an educational booklet, but our recent RCT infirmed these results.^6,7^ Despite conflicting evidence in ALBP, it is unknown whether these interventions could influence mechanistic outcomes. Understanding how interventions work in ALBP may reveal precise treatment targets, increase their effectiveness, and reduce disabilities induced by LBP.

Increased pain sensitivity was identified in LBP,^8^ with greater effect sizes in chronic LBP (CLBP) than in ALBP.^9^ Altered spinal motor control was also evidenced in LBP. The “flexion-relaxation phenomenon”,^10^ a complete relaxation of the lumbar erector spinae (LES) muscles during full trunk flexion in pain-free individuals, is absent in ∼55% of individuals with CLBP, suggesting LES overactivation.^11^ In ALBP, one study observed LES overactivation compared to pain-free individuals,^12^ which has been hypothesised as a mechanism to protect the painful posterior passive spine structures.^10^ However, the persistence of LES overactivation could contribute to maintain LBP related disability on the long-term – for example – by increasing spinal loading.^13–15^

Heatwrap produces a rapid and sustained skin and muscle temperature increase, resulting in enhanced blood flow.^16^ It has been suggested that heat therapy could accelerate metabolism, increase nutrients and oxygen to the injured tissues, reduce inflammatory markers and subsequently decrease afferent fiber sensitization.^17,18^ In addition, it was hypothesized to produce analgesia by activating thermoreceptors and engaging mechanisms such as gate control theory.^19,20^ However, these theories are limited by their reliance on observational studies in pain-free individuals and chronic wound populations.^17,18^ As for exercise, a mechanism that could explain its benefits is the “exercise-induced hypoalgesia – EIH”,^21,22^ described as an acute pain sensitivity reduction following aerobic, isometric, dynamic resistance^22^ and stretching^23^ exercises in pain-free individuals, but also in CLBP. Moreover, exercise was shown to normalize trunk muscle activation during trunk flexion in CLBP.^24–26^ However, none of these mechanisms were studied in ALBP.

With their respective mechanisms, heatwrap and exercise may contribute to normalize sensorimotor alterations in ALBP. This study aimed to (i) assess immediate and short-term effects of heatwrap alone or combined with exercise, compared to a sham heatwrap, on pain sensitivity, LES activation during trunk flexion, current pain intensity and trunk flexion ROM, and (ii) explore if changes in pain sensitivity and LES activation were associated with changes in clinical outcomes (pain intensity and disability) from baseline to 1-week follow-up. We hypothesized that (i) heatwrap and exercise would improve all outcomes compared to other interventions, and that (ii) a decrease in pain sensitivity and LES activation would be associated with clinical symptoms’ improvement from baseline to 1-week follow-up.

## Methods

### Study design

This multi-arm RCT was registered at ClinicalTrials (NCT03986047). The protocol and results of its main objectives, that were to assess the effect of heatwrap and exercise compared to heatwrap alone or a sham heatwrap on disability and pain intensity at short-, mid- and long-term in ALBP, have been published previously.^7,27^ Ethics approval was obtained from the ethics committee of the CIUSSS de la Capitale Nationale: #2019-1731 – CER CIUSSS-CN (Québec City, QC, Canada) in accordance with the Declaration of Helsinki. Written informed consent was obtained from all participants prior to study inclusion. The study was reported in accordance with the Consolidated Standards of Reporting Trials (CONSORT) guidelines.^28,29^

Two visits took place over a 7-day period at a single research center (Cirris, Québec city). At baseline (T_0_), participants completed patient-reported outcome measure (PROM) questionnaires and answered questions on sociodemographic characteristics (Canadian Minimum Dataset for LBP– CMDS),^30^ after which sensorimotor outcomes were measured. Then, participants were randomly allocated to one of three interventions: heatwrap combined with exercise, heatwrap alone, or sham heatwrap. They met a physical therapist (PT) to receive instructions on their assigned intervention and complete a 1-hour intervention session. Sensorimotor outcomes were reassessed immediately after the intervention hour (T_1_ – immediate effect). Then, participants were instructed to perform their assigned intervention daily for 7 days. A second visit took place after the intervention week (T_2_), when the sensorimotor outcomes were measured again (short-term effects), and participants completed the PROM questionnaires (**Figure 1**).

**Figure 1.**
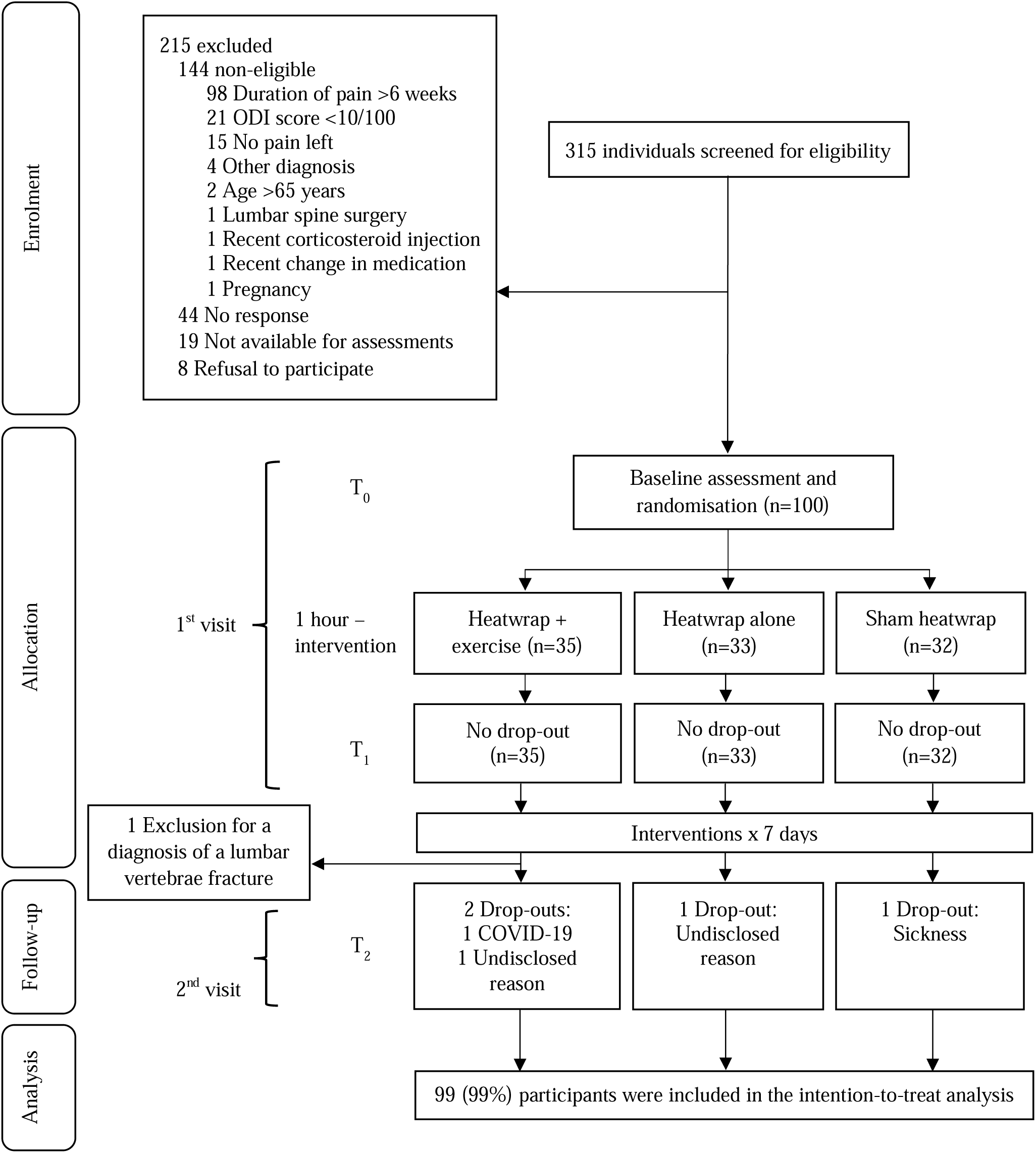
CONSORT Flow chart.

### Participants

Participants aged between 18 and 65 years with ALBP (LBP for ≤6 weeks with or without leg pain)^31^ were recruited using a convenience sampling method through the Quebec Back Pain Consortium,^32^ targeted email listings (e.g. Laval University) and Facebook ads. Participants were eligible if they scored ≥10% to the Oswestry Disability Index (ODI).^33^ They were excluded if they had experienced LBP in the three months preceding the current episode, reported symptoms of peripheral or central neural impairment (severe self-reported weakness in lower limb muscles, reduced sensitivity/ pins and needles in both lower limbs) or presented signs of a serious pathology (e.g., fracture, spinal infection, tumor). Participants were also excluded if they had a recent change in a medication that could influence pain (e.g., antidepressant, opioid, etc.), were pregnant, had a diagnosis of fibromyalgia or arthritis, had cognitive impairments, received a cortisone infiltration within the previous 6 weeks, or had a history of a lumbar spine surgery. A phone screening was done by a PT to confirm eligibility.

### Randomisation and blinding

The evaluator who measured the sensorimotor outcomes was blinded to participants’ intervention allocation, while the PT who instructed participants on the interventions was blinded to their baseline assessment results. At enrollment, participants were blinded to the specific objectives of the study and to the interventions assigned to other groups. They were solely informed that the objective of the trial was to evaluate the effects of non-pharmacological interventions in ALBP. A randomisation list was generated with a computer random number generator in a 1:1:1 allocation ratio in random blocks of 3 or 6 participants by an independent research assistant not involved in data collection or analysis. Randomisation was stratified by self-reported sex and ODI score (≤19: low or ≥20: high disability). The allocation was concealed using sealed, opaque envelopes.

### Interventions

During their one-hour session with a PT at the first visit, all participants received evidence-based advice about LBP management (i.e., staying active, avoiding bed rest, activity modification and reassurance). This information was provided in a written handout, reviewed and explained by the PT during the session, and individualized as needed based on each participant’s presentation. A heatwrap that reaches up to 40°C within 30 minutes and maintains this temperature for 8 hours^34^ (ThermaCare®, Pfizer inc., New York, USA) was applied to the lumbar region in participants assigned to both heatwrap groups, whereas a heatwrap cooled down to room temperature was used for participants in the sham heatwrap group. In addition, participants allocated to the heatwrap and exercise group received an individualised exercise program prescribed by the PT based on a physical assessment and performed the exercises during the one-hour session. Given that no specific exercise is recommended for ALBP,^35^ the decision to use individualised exercises was taken to reflect clinical reasoning and usual practice. The exercise program included 3 or 4 exercises selected from a predetermined list covering four categories: functional task, trunk muscle strengthening, lumbar spine mobility, and preferential direction when applicable. A detailed list of exercises that could be prescribed and the Consensus on Exercise Reporting Template are available in the published protocol.^27^

After the first visit, participants carried out their assigned intervention program at home for 7 days. They were asked to avoid consulting other healthcare professionals for their ALBP during this period, and to record daily adherence in a logbook (number of hours wearing the heatwrap and sets of exercises performed). Participants in both heatwrap groups were instructed to wear the heatwrap for 8 consecutive hours during the day for 7 days. Participants in the sham heatwrap group were asked to follow the same schedule to control for potential contextual, supportive and sensory effects of the heatwrap. Participants in the heatwrap and exercise group were asked to perform their exercises during 30 minutes per day on 5 days during the week.

### Outcomes – Quantitative sensory testing (QST)

Two QST measures (pressure pain threshold – PPT and temporal summation of pain – TSP) were performed by a trained PT at 2 sites: locally and remotely from the painful area.^36^ Local assessments provide information on both peripheral and central sensitization, whereas remote assessments reflect potential central sensitization.^37^ PPT is a static QST measurement reflecting the basal state of the nociceptive system.^38^ TSP is a dynamic test reflecting central pain processing.^39^ The testing sequence at all time points was randomised using a computer random number generator to minimize order effects. For familiarisation purposes, PPT and TSP were applied once over the wrist flexor muscles and on the dorsal aspect of the hand, respectively. Before each QST procedure, standardized instructions freely translated into French from the German Research Network on Neuropathic Pain were provided.^40^

PPT was assessed with a digital algometer (1cm^2^ probe–FPIX, Wagner Instruments, Greenwich, USA) at two sites: (i) on the LES muscles 2-3 cm lateral to L4-L5 on the most painful side, or on the right side if pain was bilateral, and (ii) the tibialis anterior (TA) contralateral to back PPT. Pressure was applied at a rate of 0.5kg/cm^2^/s and 3 consecutive trials were performed with a 30-second pause between measurements. Participants were tested in prone position for back PPT, and supine position for TA PPT. PPT demonstrates good to excellent within- and between-session reliability in individuals with LBP.^41,42^

TSP was assessed with a Pinprick stimulator (256 mN, MRC Systems GmbH, Heidelberg, Germany) at two sites: (i) the L4-L5 intervertebral joint line and (ii) on the dorsal aspect of the 3^rd^ cuneiform bone of the foot contralateral to the most painful side. A single stimulus was applied to the tested site, and the participant was asked to rate pain intensity on a 0 (no pain) to 10 (worst imaginable pain) numerical pain rating scale (NPRS).^43^ Immediately after, 10 stimuli were applied at a frequency of 1 Hz monitored by a flashing metronome out of the participant sight, and the participant was asked to rate the intensity of the worst pain felt during the procedure. The TSP outcome was the difference between the highest pain intensity over the 10 stimuli and the pain intensity during a single stimulus.^44^ This procedure was performed 3 consecutive times with a 30-second break between measurements. The back TSP was performed with the participant in prone position, and the foot TSP testing in supine. The within-session reliability of TSP in LBP is considered as good,^42^ and the between-session reliability in clinical populations including LBP as moderate.^45^

### Outcomes – surface electromyography and ROM

Wireless surface electromyography (sEMG) sensors (TrignoTM Wireless System, Delsys) were placed over the L5 and T12 erector spinae muscles on the most painful side (or the right side if the pain was bilateral)^46^ to record muscle activity at a sampling rate of 1000 Hz. The sensors included inertial measurement units sampling 3-dimension acceleration at 148 Hz. The skin was shaved if needed and cleaned with alcohol before sensor placement. Sensors were fixed on the skin with a double-sided tape and oriented parallel to muscle fibers. Participants were asked to stand still for 3 seconds with feet at shoulder width, bend the trunk forward to reach their full trunk flexion in 5 seconds, remain in full flexion for 3 seconds, and return to the initial position over 5 seconds (**Figure 2**). This procedure was repeated 3 times, with a 30-second interval between measurements. Participants were instructed to place one hand over the other during the trunk flexion task. At maximal flexion, the distance from the tip of the third finger to the floor was measured using a tape measure at each trial as a proxy for spine ROM. This procedure has an excellent intra-rater reliability (ICC from 0.92 to 0.97) for forward flexion of the trunk.^47^

**Figure 2.**
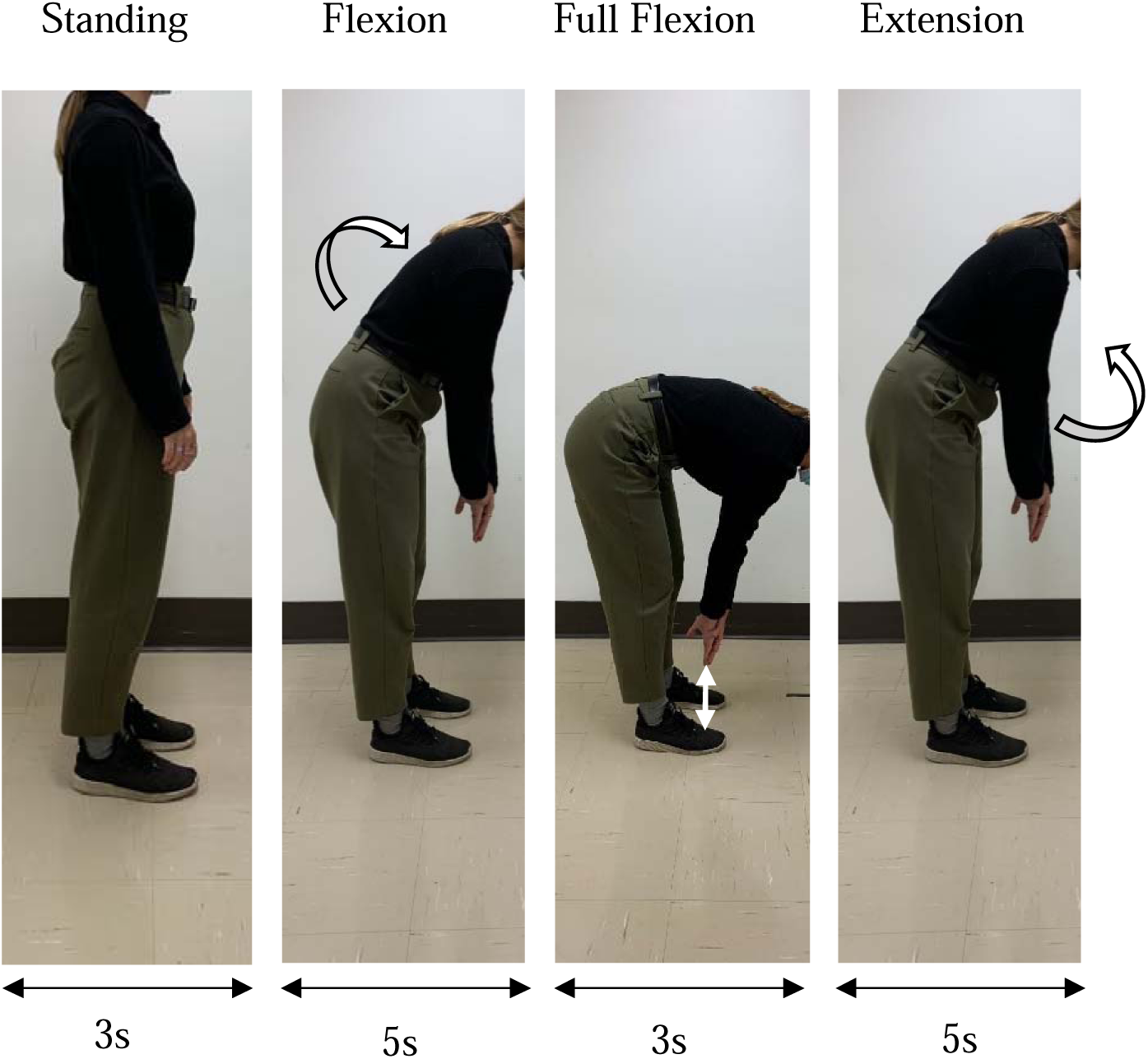
Trunk flexion task for sEMG recording.

Raw sEMG data from both sensors were imported into MATLAB R2019a (The Mathworks inc., Natick, Massachusetts, USA), full-wave rectified and band-pass filtered with a 4^th^ order Butterworth filter (20-450 Hz). Raw sagittal plane acceleration data were imported into MATLAB and resampled to fit sEMG sampling rate (1000 Hz). A moving average with a 250 ms sliding window was used to visualize the sEMG signal, and a custom graphical user interface allowed to overlay the raw acceleration signal to the rectified and filtered sEMG signal. This interface was used to identify each phase of the trunk flexion: (i) standing, (ii) flexion, (iii) full flexion, and (iv) extension. The peak sEMG signal was then identified at each phase and averaged through the 3 trials. The mean peak values of the full flexion and extension phases were used to calculate the flexion-relaxation ratios (FRR) at each sensor (L5 and T12), as recommended in a meta-analysis by Gouteron et al.^11^ FRR has moderate to high test-retest reliability.^48^

### Patient reported outcome measures

Participants rated their current pain intensity using the NPRS at T_0_, T_1_ and T_2_ in a standardized seated position. Average LBP intensity over the last 7 days (NPRS) and pain-related disability (ODI) were self-rated by participants at T_0_ and T_2_. Central sensitization symptoms (Central sensitization inventory),^49^ pain-related fear (17-item Tampa Scale for Kinesiophobia questionnaire)^50^ and pain catastrophizing (Pain Catastrophizing Scale)^51^ were assessed at baseline to characterise the study participants and used as covariates in the statistical models.

### Sample size

The sample size was calculated using G*Power 3.1.9.7 based on between-group differences in the primary outcome (ODI) of the main RCT.^7,27^ Thus, the sample size was not specifically powered for the sensorimotor outcomes. However, sensitivity analyses were performed to estimate the effect sizes that could be detected given the available sample size. Considering an α=0.05, statistical power (1-β) of 0.80 and group sizes ranging from n=32 to n=34, the minimum detectable effect size was moderate (Cohen’s d=0.61-0.62).

### Statistical analysis

Statistical analyses were realised using SPSS statistics v.30.0. Baseline demographic characteristics and PROMs were presented as counts and percentages for categorical variables and mean and standard deviations (SD) for continuous variables. Normality and variance homogeneity were verified by examining residuals’ distribution, skewness and kurtosis, and computing the Shapiro-Wilk test. As the results for the effect of the interventions on PROMs were previously published,^7^ no analysis on the effect of the interventions on these outcomes is presented. Within-sample mean differences (MD) between baseline and the 1-week follow-up were reported for pain-related disability and average pain-intensity over the last 7 days for the regression analyses (objective [ii] – see next section).

For objective (i), linear mixed models (LMMs) were used to compare interventions’ efficacy on pain sensitivity (TS and PPT), sEMG ratios, current pain intensity and trunk flexion ROM with intention-to-treat analyses and a random effect for participants’ intercept. LMMs use a restricted maximum likelihood estimation approach assuming data to be missing at random, allowing all available data to be included.^52^ The models included terms for Group and Time (T_0_, T_1_ and T_2_), Group x Time interaction, and as recommended, stratification variables (sex and ODI [≤19 and ≥20]), and a prognostic factor (age)^53^ as covariates.^54,55^ For objective (ii), linear regression equations were computed to assess the associations between the changes in TSP, PPT and FRRs (independent variables) and the changes in pain intensity over the last 7 days and disability (dependent variables) from baseline to 1-week follow-up. Pain sensitivity models (TSP and PPT) were adjusted for age, sex, and baseline Pain catastrophizing scale and Central sensitization inventory scores. FRR models were adjusted for age, sex, and baseline Pain catastrophizing scale and Tampa scale for kinesiophobia scores. Potential confounders were chosen based on their hypothesized associations with both independent and dependent variables.^56^ The α level was set at 0.05 for all analyses.

## Results

From April 2019 to March 2023, 315 participants were screened for eligibility, and 100 were enrolled and allocated to an intervention group. One participant in the heatwrap and exercise group was excluded retrospectively due to a confirmed lumbar vertebrae fracture that likely preceded study enrollment. Thus, data from 99 participants were included in the analyses (**Figure 1**). Baseline demographic and clinical characteristics for each group are presented in **Table 1**. Most participants reported female sex (62%), the mean age was 36.4 years (SD 12.8), the average pain intensity over the last week was 5.1/10 (SD 1.9), and the mean disability level assessed with ODI was 22.8% (SD 10.9).

**Table 1.** Baseline demographic and clinical data.

|  | Heatwrap and exercise (n=34) | Heatwrap alone (n=33) | Sham heatwrap (n=32) | Total (n=99) |
| --- | --- | --- | --- | --- |
| **Demographic characteristics** |  |  |  |  |
| Female, n (%) | 20 (58.8) | 21 (63.6) | 20 (62.5) | 61 (62) |
| Age (years), mean [SD] | 35.1 [12.1] | 36.9 [13.6] | 37.3 [13.0] | 36.4 [12.8] |
| BMI (kg/m<sup>2</sup>), mean [SD] | 25.3 [3.7] | 27.1 [5.5] | 27.6 [5.8] | 26.7 [5.1] |
| Ethnicity, n (%) |  |  |  |  |
| Metis (Indigenous people in Canada) | 1 (3) | 0 (0) | 0 (0) | 1 (1) |
| White | 22 (65) | 24 (73) | 28 (88) | 74 (75) |
| Chinese | 3 (9) | 0 (0) | 0 (0) | 3 (3) |
| Black | 2 (6) | 4 (12) | 1 (3) | 7 (7) |
| Latin American | 3 (9) | 2 (6) | 0 (0) | 5 (5) |
| Arabic | 4 (12) | 4 (12) | 3 (9) | 11 (11) |
| South-East Asian | 0 (0) | 0 (0) | 1 (3) | 1 (1) |
| Occidental Asian | 0 (0) | 1 (3) | 0 (0) | 1 (1) |
| **Clinical outcomes** |  |  |  |  |
| Participants with history of LBP, n (%) | 24 (70.6) | 20 (60.6) | 19 (59.4) | 63 (64) |
| LBP duration (days), mean [SD] | 24.2 [12.2] | 26.2 [11.2] | 25.4 [12.4] | 25.2 [11.9] |
| Current pain intensity (NPRS), mean [SD] | 2.9 [1.9] | 2.9 [2.4] | 2.9 [1.9] | 2.9 [2.1] |
| Average pain intensity over the last week (NPRS), mean [SD] | 4.7 [1.9] | 5.5 [1.9] | 5.2 [1.7] | 5.1 [1.9] |
| Disability (ODI), mean [SD] | 22.9 [10.6] | 23.2 [10.5] | 22.4 [11.9] | 22.8 [10.9] |
| Central sensitization symptoms (CSI), mean [SD] | 28.9 [13.9] | 32.1 [16.4] | 27.2 [10.0] | 29.4 [13.7] |
| Pain-related fear (TSK), mean [SD] | 36.7 [7.4] | 35.6 [6.3] | 36.7 [6.9] | 36.3 [6.8] |
| Pain catastrophizing (PCS), mean [SD] | 12.9 [9.6] | 16.5 [9.7] | 14.3 [8.9] | 14.6 [9.4] |
| SD, standard deviation; BMI, body mass index; LBP, low back pain; NPRS, numerical pain rating scale; ODI, Oswestry Disability Index; CSI, central sensitization inventory; TSK, Tampa Scale for Kinesiophobia; PCS, Pain Catastrophizing Scale. |  |  |  |  |

### Effect of interventions on sensorimotor outcomes

No analyse for objective (i) showed a significant Group x Time interaction (**Table 2**, **Figure 3**). A significant effect of Time was observed for lumbar (F=6.22 p<0.002) and tibialis anterior (F=7.53, p<0.001) PPT, for lumbar (F=9,87, p<0.001) and foot (F=9.03, p<0.001) TSP, for trunk flexion ROM (F=17.42, p<0.001) and for current pain intensity (F=24.35, p<0.001). No significant effect of Time was detected for FRR at L5 and T12 (p>0.26).

**Table 2.** Between-group estimated mean differences at each time-point for each outcome and 95% confidence intervals (95% CI)

|  | Mean difference (95% CI)<sup>a</sup> |  |  |
| --- | --- | --- | --- |
|  | Heatwrap and exercise vs sham (heatwrap & exercise minus sham) | Heatwrap alone vs sham (heatwrap minus sham) | Heatwrap and exercise vs heatwrap alone (heatwrap & exercise minus heatwrap alone) |
| Lumbar PPT (kg/cm<sup>2</sup>) |  |  |  |
| T<sub>0</sub> | -0.25 (-1.37 to 0.87) | -0.04 (-1.16 to 1.09) | -0.21 (-1.32 to 0.90) |
| T<sub>1</sub> | -0.52 (-1.64 to 0.60) | -0.02 (-1.14 to 1.11) | -0.51 (-1.61 to 0.60) |
| T<sub>2</sub> | -0.40 (-1.52 to 0.73) | 0.24 (-0.89 to 1.37) | -0.64 (-1.75 to 0.48) |
| Tibialis anterior PPT (kg/cm<sup>2</sup>) |  |  |  |
| T<sub>0</sub> | -0.19 (-1.35 to 0.96) | 0.05 (-1.11 to 1.21) | -0.24 (-1.39 to 0.90) |
| T<sub>1</sub> | 0.10 (-1.06 to 1.25) | 0.28 (-0.88 to 1.44) | -0.18 (-1.33 to 0.96) |
| T<sub>2</sub> | 0.11 (-1.06 to 1.27) | 0.11 (-1.06 to 1.27) | 0.00 (-1.15 to 1.15) |
| Lumbar TSP (NPRS) |  |  |  |
| T<sub>0</sub> | 0.35 (-0.33 to 1.03) | -0.07 (-0.75 to 0.62) | 0.42 (-0.25 to 1.09) |
| T<sub>1</sub> | 0.49 (-0.19 to 1.17) | -0.40 (-1.08 to 0.29) | 0.89 (0.22 to 1.56) |
| T<sub>2</sub> | 0.41 (-0.28 to 1.10) | -0.35 (-1.04 to 0.35) | 0.76 (0.08 to 1.44) |
| Foot TSP (NPRS) |  |  |  |
| T<sub>0</sub> | 0.48 (-0.20 to 1.17) | 0.02 (-0.67 to 0.71) | 0.46 (-0.21 to 1.14) |
| T<sub>1</sub> | 0.74 (0.05 to 1.42) | -0.15 (-0.83 to 0.54) | 0.88 (0.21 to 1.55) |
| T<sub>2</sub> | 0.41 (-0.29 to 1.10) | -0.40 (-1.10 to 0.29) | 0.81 (0.13 to 1.49) |
| FRR L5 |  |  |  |
| T<sub>0</sub> | -0.02 (-0.15 to 0.11) | -0.06 (-0.18 to 0.07) | 0.03 (-0.09 to 0.16) |
| T<sub>1</sub> | -0.04 (-0.17 to 0.09) | -0.07 (-0.20 to 0.06) | 0.03 (-0.10 to 0.15) |
| T<sub>2</sub> | -0.09 (-0.22 to 0.04) | -0.07 (-0.20 to 0.06) | -0.02 (-0.15 to 0.11) |
| FRR T12 |  |  |  |
| T<sub>0</sub> | -0.05 (-0.16 to 0.07) | 0.02 (-0.10 to 0.13) | -0.07 (-0.18 to 0.05) |
| T<sub>1</sub> | -0.07 (-0.18 to 0.05) | 0.001 (-0.12 to 0.12) | -0.07 (-0.18 to 0.05) |
| T<sub>2</sub> | -0.10 (-0.21 to 0.02) | -0.03 (-0.15 to 0.09) | -0.07 (-0.18 to 0.05) |
| ROM (cm) |  |  |  |
| T<sub>0</sub> | -4.10 (-9.77 to 1.56) | -1.73 (-7.42 to 3.96) | -2.37 (-7.98 to 3.24) |
| T<sub>1</sub> | -3.33 (-9.00 to 2.34) | -0.68 (-6.37 to 5.01) | -2.65 (-8.27 to 2.97) |
| T<sub>2</sub> | -4.04 (-9.72 to 1.65) | -0.68 (-6.38 to 5.02) | -3.36 (-8.99 to 2.27) |
| Current pain intensity (NPRS) |  |  |  |
| T<sub>0</sub> | -0.04 (-0.93 to 0.85) | 0.02 (-0.88 to 0.92) | -0.06 (-0.95 to 0.82) |
| T<sub>1</sub> | -0.11 (-1.00 to 0.78) | -0.16 (-1.06 to 0.73) | 0.06 (-0.83 to 0.94) |
| T<sub>2</sub> | 0.37 (-0.54 to 1.27) | 0.26 (-0.65 to 1.17) | 0.10 (-0.80 to 1.00) |
| PPT, pressure pain threshold; TSP, temporal summation of pain; NPRS, numerical pain rating scale; FRR, flexion-relaxation ratio; ROM, range of motion (distance from 3<sup>rd</sup> finger to floor during full trunk flexion).<br><sup>a</sup>Mean differences are reported as the subtraction of the mean of the second group from that of the first group (e.g., the first column presents the mean difference of the heatwrap and exercise group minus the sham. A negative value for PPT means individuals in the heatwrap + exercise were more sensitive compared to sham). |  |  |  |

**Figure 3.**
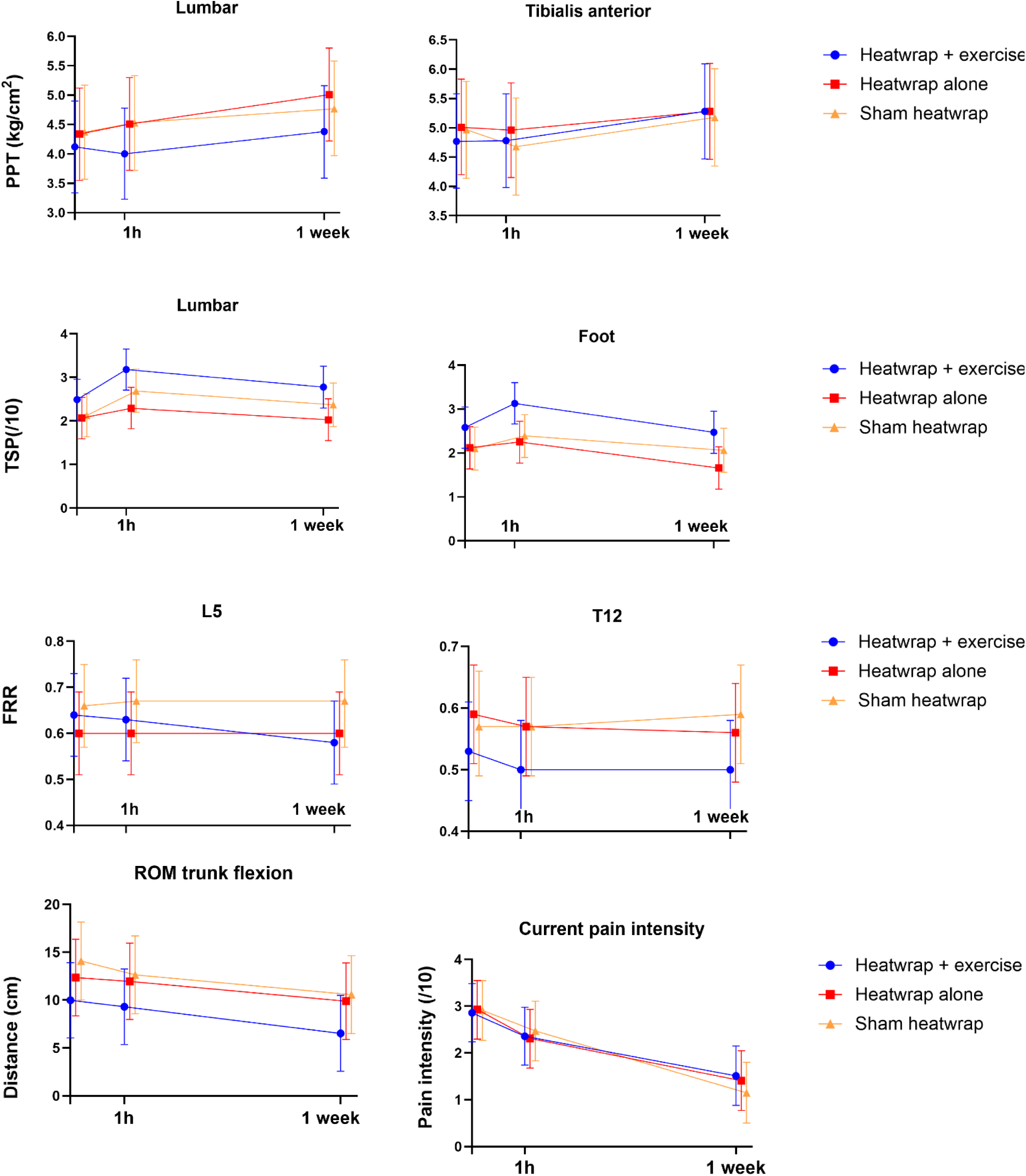
Mean and 95% confidence interval of pain sensitivity, flexion-relaxation ratios, current pain intensity, and amplitude of global trunk flexion for each group at each time-points. PPT, pressure pain threshold; TSP, temporal summation of pain; FRR, flexion-relaxation ratio; ROM, range of motion.

Pairwise comparisons – between timepoints and regardless of groups – showed that PPT measures increased at T_2_ (one week) compared to both T_1_ (one hour) and T_0_ (baseline), reflecting reduced pain sensitivity. Also, both TSP measures increased at T_1_ compared to T_0_, reflecting increased pain sensitivity, and decreased at T_2_ compared to T_1_, but not compared to T_0,_ suggesting a return to baseline values. The finger-to-floor distance during full trunk flexion decreased at T_2_ compared to both T_1_ and T_0_, depicting increased trunk ROM. Current pain intensity significantly decreased at T_1_ compared to T_0_, and further decreased after a week (T_2_) compared to both T_1_ and T_0_. Within-group mean differences are presented in **Table 3**. **Supplementary Table 1** displays the means for each group and the complete sample for all outcomes at each time-point.

**Table 3.** Within-sample estimated mean differences and 95% CI of sensorimotor and clinical outcomes at all time-points (time effect).

| Outcomes | Mean difference (95% CI) |  |  |
| --- | --- | --- | --- |
| | $\Delta 1h$-baseline | $\Delta 1week$-baseline | $\Delta 1week-1h$ |
| **Clinical** |  |  |  |
| Pain-related disability (ODI) | NA | -7.8<sup>a</sup> (-9.7 to -5.9) | NA |
| Pain intensity over the last 7 days (NPRS) | NA | -2.2<sup>a</sup> (-2.6 to -1.7) | NA |
| **Sensorimotor** |  |  |  |
| Lumbar PPT (kg/cm<sup>2</sup>) | 0.07 (-0.11 to 0.25) | 0.44** (0.19 to 0.69) | 0.37* (0.12 to 0.62) |
| Tibialis anterior PPT (kg/cm<sup>2</sup>) | -0.11 (-0.33 to 0.11) | 0.33* (0.03 to 0.63) | 0.44** (0.22 to 0.67) |
| Lumbar TSP (NPRS) | 0.49** (0.26 to 0.73) | 0.17 (-0.13 to 0.46) | -0.33* (-0.57 to -0.09) |
| Foot TSP (NPRS) | 0.32* (0.06 to 0.58) | -0.20 (-0.52 to 0.12) | -0.52** (-0.78 to -0.26) |
| L5 FRR | 0.00 (-0.03 to 0.03) | -0.02 (-0.06 to 0.02) | -0.02 (-0.05 to 0.01) |
| T12 FRR | -0.02 (-0.05 to 0.01) | -0.01 (-0.05 to 0.02) | 0.01 (-0.03 to 0.04) |
| Flexion ROM (cm) | -0.84 (-1.73 to 0.04) | -3.15** (-4.27 to -2.02) | -2.31** (-3.20 to -1.41) |
| Current pain intensity (NPRS) | -0.52* (-0.90 to -0.14) | -1.54** (-1.99 to -1.09) | -1.02** (-1.41 to -0.63) |
| T<sub>0</sub>, baseline; T<sub>1</sub>, 1-hour follow-up; T<sub>2</sub>, 1-week follow-up; $\Delta 1h$-baseline, outcome measured at 1 hour minus baseline; $\Delta 1week$-baseline, outcome measured at 1 week minus baseline; $\Delta 1week-1h$, outcome measured at 1 week minus 1 hour; ODI, Oswestry Disability Index; NA, not assessed; NPRS, numerical pain rating scale; PPT, pressure pain threshold; TSP, temporal summation of pain; FRR, flexion-relaxation ratio; ROM, range of motion.<br><sup>a</sup>Scores were significantly different at 1 week compared to baseline (see Côté-Picard et al., 2026)<br>*p<0.05; **p<0.001. |  |  |  |

### Association estimates of changes in sensorimotor and clinical outcomes at 1-week compared to baseline

The results of the linear regressions are presented in **Table 4**. None of the 12 models showed statistically significant associations between changes in sensorimotor outcomes and changes in clinical outcomes from baseline to 1-week.

**Table 4.**
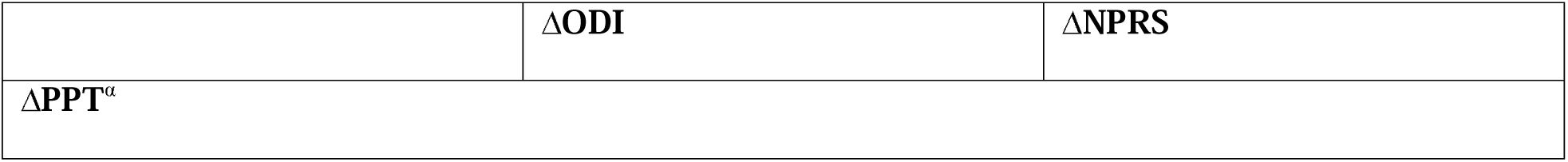
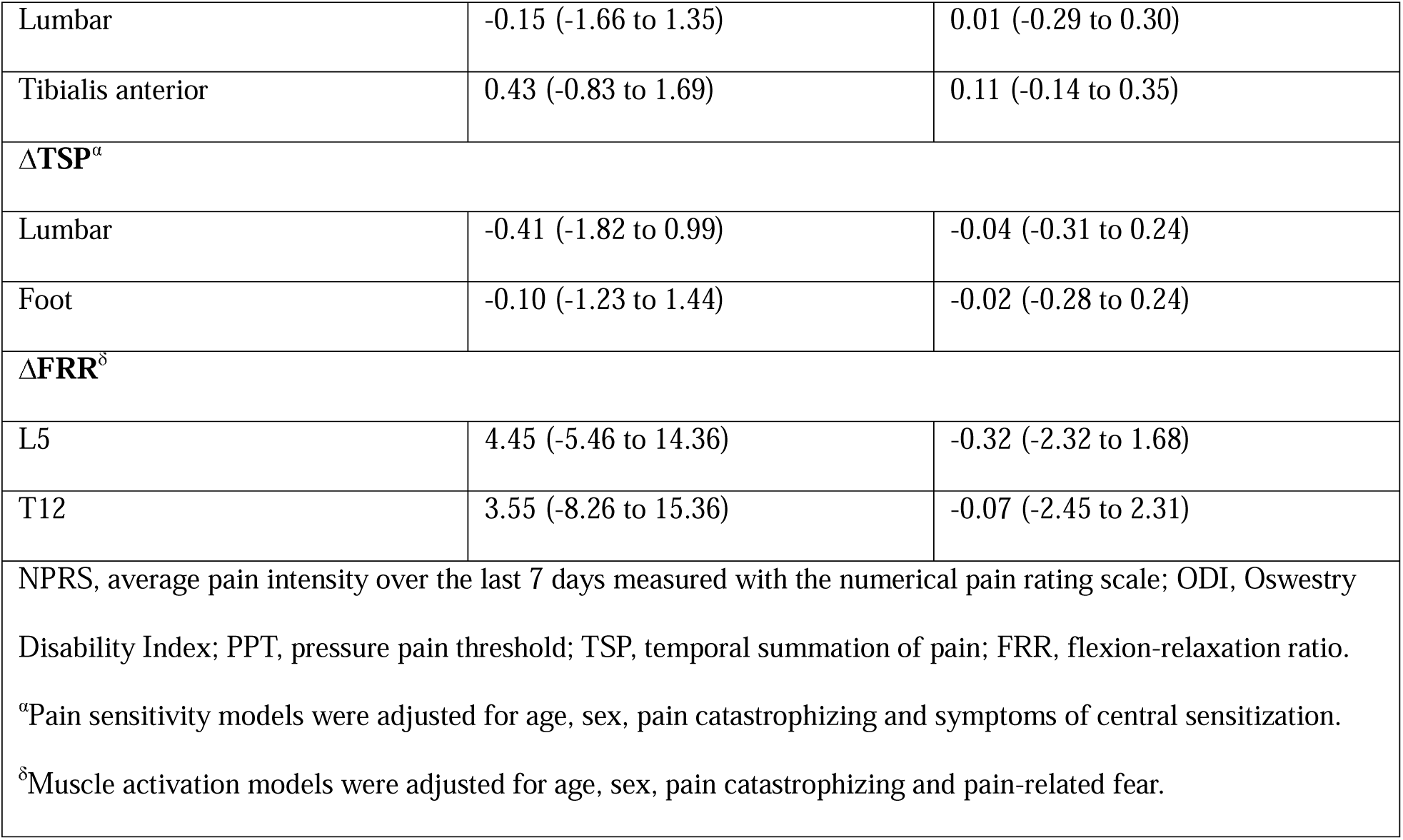
Linear regressions (unstandardized β coefficient [95% confidence interval]) between sensorimotor and clinical changes from baseline to 1-week follow-up.

| | $\Delta ODI$ | $\Delta NPRS$ |
| --- | --- | --- |
| $\Delta PPT^a$ | | |
| Lumbar | -0.15 (-1.66 to 1.35) | 0.01 (-0.29 to 0.30) |
| Tibialis anterior | 0.43 (-0.83 to 1.69) | 0.11 (-0.14 to 0.35) |
| $\Delta\text{TSP}^a$ | | |
| Lumbar | -0.41 (-1.82 to 0.99) | -0.04 (-0.31 to 0.24) |
| Foot | -0.10 (-1.23 to 1.44) | -0.02 (-0.28 to 0.24) |
| $\Delta\text{FRR}^\delta$ | | |
| L5 | 4.45 (-5.46 to 14.36) | -0.32 (-2.32 to 1.68) |
| T12 | 3.55 (-8.26 to 15.36) | -0.07 (-2.45 to 2.31) |
| NPRS, average pain intensity over the last 7 days measured with the numerical pain rating scale; ODI, Oswestry Disability Index; PPT, pressure pain threshold; TSP, temporal summation of pain; FRR, flexion-relaxation ratio.<br><sup>a</sup>Pain sensitivity models were adjusted for age, sex, pain catastrophizing and symptoms of central sensitization.<br><sup>δ</sup>Muscle activation models were adjusted for age, sex, pain catastrophizing and pain-related fear. |  |  |

## Discussion

To our knowledge, this is the first RCT to investigate the mechanisms of heatwrap alone or combined with exercise in ALBP. For the first objective, we hypothesised that heatwrap combined with exercise would have a greater impact on pain sensitivity, erector spinae muscle activation during trunk flexion, current pain intensity and trunk flexion ROM, than heatwrap alone or a sham heatwrap. However, the results infirm this hypothesis as no significant differences between the intervention groups were found for any outcomes. Nonetheless, significant time effects were observed for PPT, TSP, trunk flexion ROM and current pain intensity. Specifically, at one-week follow-up compared with both baseline and the one-hour assessment, local and remote PPT increased, trunk flexion ROM increased, and current pain intensity decreased. An increase in TSP was observed locally and remotely at one hour compared with baseline. We believe this TSP change may be explained by transient skin sensitization induced by the high number of stimuli administered within a single session (three blocks of 10 stimuli performed twice [pre–post intervention]), as previously reported in the literature.^42^ This increase in TSP was no longer present at the one-week follow-up. Besides, no associations were found between changes in pain sensitivity or muscle activation and clinical symptoms at one week compared to baseline, thereby not supporting the hypothesis for the second objective.

### Pain sensitivity outcomes

The lack of between-group differences across all sensorimotor outcomes is consistent with the results of the main RCT, where no intervention was significantly superior to reduce pain-related disability and pain intensity.^7^ The absence of an immediate (within-session) effect of heatwrap combined with exercise on pain sensitivity, despite evidence suggesting EIH in CLBP,^21,22^ could be due to the fact that only a limited number of isometric exercises (e.g. abdominal plank), and no aerobic exercises could be prescribed, both of which have been shown to elicit EIH in CLBP.^21,22^ Besides, the lack of between-group effect of both heatwrap groups on sensory outcomes, compared to a sham heatwrap, suggests that it did not induce larger hypoalgesia. Moreover, the increase in local and remote PPT at one week – and not at 1 hour – in all groups suggests that (i) none of the group induced immediate hypoalgesia and (ii) the heatwrap interventions had no short-term effects beyond that of the sham heatwrap, or the effects were so small that they could not be captured given the study’s sample size. This finding also suggests that the tactile stimulus of sham heatwrap (and real heatwrap) might have been sufficient to foster pain modulation in activating the gate-control theory. Other possibilities are that the observed increase in PPT reflects either the natural evolution of pain sensitivity in ALBP and/or the effect of standardised advice provided to all participants. An RCT reported that education-based interventions increased PPT in individuals with knee osteoarthritis.^57^ However, we cannot confirm this hypothesis as we did not have a group without advice. The results also suggest that a normalization in pain sensitivity occurred with symptoms’ improvement, although they were not significantly associated at the one-week follow-up.

The absence of association between pain sensitivity changes and clinical improvement suggests that in ALBP, short-term changes in pain sensitivity may not directly drive changes in pain and disability. This result is however partly consistent with findings from previous studies. In a pilot work by Massé-Alarie et al.,^58^ changes in both PPT and TSP were significantly associated with changes in disability, but not pain intensity, at a 6-week follow-up in participants with CLBP.^58^ Similarly, the results from Nim et al. (2021) showed no association between changes in pain sensitivity and clinical improvement in patients with CLBP.^59^

### Erector spinae muscle activation outcomes

The lack of a between-group effect on FRR outcomes suggests that the interventions did not reduce LES activation in full flexion. This does not support our initial hypothesis and the theories on the effects of heatwrap and exercises on muscle activation. To our knowledge, no other study examined the effect of heat therapy on FRR or on objective muscle stiffness measures in participants with LBP. However, a systematic review by Bleakley et al., (2013) assessed the effect of thermal agents on soft tissues’ properties and found similar results as heat therapy had no effect on muscle stiffness.^20^ The absence of an effect of exercise on muscle activation outcomes contrasts with the results of many studies in which exercise normalized the FRR in participants with CLBP.^24–26,60^ An explanation is that the FRR might not have been altered, in average, in our participants’ sample, impeding on the possibility that interventions have an effect on this outcome.

Given that current pain intensity decreased for all groups at both one hour and one week and that trunk flexion ROM and clinical symptoms improved at one week compared to baseline,^7^ it was surprising that no time effect was observed for erector spinae muscle activation. Considering the theories on motor control alterations in LBP,^13^ a decrease in FRR should have occurred with symptoms’ improvements. This impeded the possibility that changes in FRR outcomes and symptoms at one week be associated, thus infirming our initial hypothesis. This result partly contrasts from a systematic review by Wernli et al. (2020) who found low quality evidence that changes in lumbar movement (including FRR) were associated with changes in pain or disability 31% of the time in LBP.^15^ However, our results align with those of Shahvarpour et al. (2017),^61^ in which no change in FRR was observed after an 8-week stabilization exercise program in participants with subacute/chronic LBP, even though symptoms significantly improved.^61^ Thus, there remains uncertainty regarding the relationship between these variables. Our team previously published a study in which the participants of the present RCT were grouped based on their sensory, motor and psychological characteristics,^62^ and found two groups with predominant motor alteration features (sensorimotor and psychomotor groups.) We may suppose that the interventions had a greater effect on FRR or that an improvement in FRR might have been associated with clinical improvements in these groups, but this remains to be tested.

### Limitations

One might question if the duration of interventions was sufficient to elicit measurable effects on sensorimotor outcomes. In a previous study that assessed the effect of heatwrap, a rapid and sustained increase in skin and muscle temperature was shown; however, muscle temperature began to increase only after 30 minutes of application and continued to increase until the end of treatment (2 hours).^16^ It is possible that the one-hour application was insufficient for heatwrap to produce detectable effects on pain sensitivity and muscle activation. However, exercises should have provided an acute effect on sensorimotor outcomes as shown in other studies,^22^ but as mentioned, the types of exercises provided in the current RCT might not have been appropriate to produce such effects. Moreover, it is possible that, on average, participants did not present with altered FRR. A limitation that could explain this is that the recruitment took place in the community, as opposed to clinical settings in other studies,^12,60^ so the participants’ condition might not have been severe enough for motor alterations to occur. Also, it is questionable whether FRR was an adequate proxy of muscle stiffness or spasms, as a potential target for heat and exercise therapy. Indeed, FRR may be influenced by many factors including pain-related fear of movement^63^ and trunk flexion ROM,^64^ which, if not complete, might not allow muscles to relax. Other measures of muscle stiffness might be better suited to assess the mechanisms of heatwrap on muscle properties. Finally, our sample size allowed to capture moderate to high effect sizes. Thus, it was under-powered to capture smaller effects, leading to potential type II errors.

### Conclusions

In this RCT, heatwrap alone or combined with exercise had no significant immediate and short-term effect on pain sensitivity, LES muscle activity, trunk flexion ROM and current pain intensity in ALBP compared to a sham heatwrap. Accordingly, none of the hypothesised mechanisms underlying the effects of heatwrap and exercise could be supported. Also, changes in sensorimotor outcomes were not associated with changes in clinical outcomes at one week compared to baseline. Future studies assessing the mechanisms of heatwrap and exercises in ALBP must be conducted while recruiting participants with potentially more altered sensorimotor features such as in clinical settings and using different proxies that may more accurately capture muscle stiffness and related neuromuscular properties.

## Data Availability

All data produced in the present study are available upon reasonable request to the authors.

## Acknowledgments

We acknowledge the help of Valérie Jomphe, statistician, for statistical analyses, and of Alexandre Desgagné-Lebeuf, engineer, for the treatment of sEMG data. We also thank all participants recruited in this study.

## Abbreviations

ALBP: Acute low back pain
CLBP: Chronic low back pain
CNS: Central nervous system
FRR: Flexion relaxation ratio
ICC: Intraclass correlation coefficient
LBP: Low back pain
LES: Lumbar erector spinae
MD: Mean difference
NPRS: Numerical pain rating scale
ODI: Oswestry Disability Index
PPT: Pressure pain threshold
PROMs: Patient-reported outcome measure
PT: Physical therapist
QST: Quantitative sensory testing
RCT: Randomized controlled trial
ROM: Range of motion
SD: Standard deviations
sEMG: Surface Electromyography
TA: Tibialis anterior
TSP: Temporal summation of pain

## REFERENCES

1. Ferreira ML, de Luca K, Haile LM, et al. Global, regional, and national burden of low back pain, 1990&#x2013;2020, its attributable risk factors, and projections to 2050: a systematic analysis of the Global Burden of Disease Study 2021. The Lancet Rheumatology. 2023;5(6):e316–e329.

2. Wallwork SB, Braithwaite FA, O’Keeffe M, et al. The clinical course of acute, subacute and persistent low back pain: a systematic review and meta-analysis. CMAJ. 2024;196(2):E29–E46.

3. Oliveira CB, Koes BW, Pinto RZ, et al. Towards global clinical practice guidelines for the management of non-specific low back pain in primary care: a review of current guideline recommendations and how they have changed over the last 30 years. The Lancet Rheumatology. 2026;8(6):e470–e485.

4. Qaseem A, Wilt TJ, McLean RM, Forciea MA, Clinical Guidelines Committee of the American College of P. Noninvasive Treatments for Acute, Subacute, and Chronic Low Back Pain: A Clinical Practice Guideline From the American College of Physicians. Ann Intern Med. 2017;166(7):514–530.

5. Gianola S, Bargeri S, Del Castillo G, et al. Effectiveness of treatments for acute and subacute mechanical non-specific low back pain: a systematic review with network meta-analysis. Br J Sports Med. 2022;56(1):41–50.

6. Mayer JM, Ralph L, Look M, et al. Treating acute low back pain with continuous low-level heat wrap therapy and/or exercise: a randomized controlled trial. Spine J. 2005;5(4):395–403.

7. Côté-Picard C, Tittley J, Mailloux C, et al. Heatwrap and exercise in acute low back pain: a multi-arm randomised controlled trial. Musculoskeletal Science & Practice. 2026;84.

8. den Bandt HL, Paulis WD, Beckwee D, Ickmans K, Nijs J, Voogt L. Pain Mechanisms in Low Back Pain: A Systematic Review With Meta-analysis of Mechanical Quantitative Sensory Testing Outcomes in People With Nonspecific Low Back Pain. J Orthop Sports Phys Ther. 2019;49(10):698–715.

9. den Bandt HL, Ickmans K, Leemans L, Nijs J, Voogt L. Differences in Quantitative Sensory Testing Outcomes Between Patients With Low Back Pain in Primary Care and Pain-free Controls. Clin J Pain. 2022;38(6):381–387.

10. Colloca CJ, Hinrichs RN. The Biomechanical and Clinical Significance of the Lumbar Erector Spinae Flexion-Relaxation Phenomenon: A Review of Literature. Journal of Manipulative and Physiological Therapeutics. 2005;28(8):623–631.

11. Gouteron A, Tabard-Fougere A, Bourredjem A, Casillas JM, Armand S, Genevay S. The flexion relaxation phenomenon in nonspecific chronic low back pain: prevalence, reproducibility and flexion-extension ratios. A systematic review and meta-analysis. Eur Spine J. 2022;31(1):136–151.

12. Qiao J, Zhang SL, Zhang J, Feng D. A study on the paraspinal muscle surface electromyography in acute nonspecific lower back pain. Medicine (Baltimore). 2019;98(34):e16904.

13. Hodges PW, Tucker K. Moving differently in pain: a new theory to explain the adaptation to pain. Pain. 2011;152(3 Suppl):S90–S98.

14. Gouteron A, Moissenet F, Tabard-Fougère A, et al. Relationship between the flexion relaxation phenomenon and kinematics of the multi-segmental spine in nonspecific chronic low back pain patients. Sci Rep. 2024;14(1):24335.

15. Wernli K, Tan JS, O’Sullivan P, Smith A, Campbell A, Kent P. Does Movement Change When Low Back Pain Changes? A Systematic Review. J Orthop Sports Phys Ther. 2020;50(12):664–670.

16. Petrofsky JS, Laymon M, Berk L, Bains G. Effect of ThermaCare HeatWraps and Icy Hot Cream/Patches on Skin and Quadriceps Muscle Temperature and Blood Flow. J Chiropr Med. 2016;15(1):9–18.

17. Wright A, Sluka KA. Nonpharmacological treatments for musculoskeletal pain. Clin J Pain. 2001;17(1):33–46.

18. Malanga GA, Yan N, Stark J. Mechanisms and efficacy of heat and cold therapies for musculoskeletal injury. Postgrad Med. 2015;127(1):57–65.

19. Trowbridge CA, Draper DO, Feland JB, Jutte LS, Eggett DL. Paraspinal musculature and skin temperature changes: comparing the Thermacare HeatWrap, the Johnson & Johnson Back Plaster, and the ABC Warme-Pflaster. J Orthop Sports Phys Ther. 2004;34(9):549–558.

20. Bleakley CM, Costello JT. Do Thermal Agents Affect Range of Movement and Mechanical Properties in Soft Tissues? A Systematic Review. Archives of Physical Medicine and Rehabilitation. 2013;94(1):149–163.

21. Patricio P, Mailloux C, Wideman TH, et al. Assessment of exercise-induced hypoalgesia in chronic low back pain and potential associations with psychological factors and central sensitization symptoms: A case-control study. Pain Pract. 2023;23(3):264–276.

22. Naugle KM, Fillingim RB, Riley JL, 3rd. A meta-analytic review of the hypoalgesic effects of exercise. J Pain. 2012;13(12):1139–1150.

23. Larouche MC, Camire Bernier S, Racine R, et al. Stretch-induced hypoalgesia: a pilot study. Scand J Pain. 2020;20(4):837–845.

24. Cho SH, Park SY. Immediate effects of isometric trunk stabilization exercises with suspension device on flexion extension ratio and strength in chronic low back pain patientss. J Back Musculoskelet Rehabil. 2019;32(3):431–436.

25. Marshall P, Murphy B. Changes in the flexion relaxation response following an exercise intervention. Spine (Phila Pa 1976). 2006;31(23):E877–883.

26. Marshall PW, Murphy BA. Evaluation of functional and neuromuscular changes after exercise rehabilitation for low back pain using a Swiss ball: a pilot study. J Manipulative Physiol Ther. 2006;29(7):550–560.

27. Cote-Picard C, Tittley J, Mailloux C, et al. Effect of thermal therapy and exercises on acute low back pain: a protocol for a randomized controlled trial. BMC Musculoskelet Disord. 2020;21(1):814.

28. Juszczak E, Altman DG, Hopewell S, Schulz K. Reporting of Multi-Arm Parallel-Group Randomized Trials: Extension of the CONSORT 2010 Statement. JAMA. 2019;321(16):1610–1620.

29. Moher D, Hopewell S, Schulz KF, et al. CONSORT 2010 explanation and elaboration: updated guidelines for reporting parallel group randomised trials. BMJ. 2010;340:c869.

30. Lacasse A, Roy JS, Parent AJ, et al. The Canadian minimum dataset for chronic low back pain research: a cross-cultural adaptation of the National Institutes of Health Task Force Research Standards. CMAJ Open. 2017;5(1):E237–E248.

31. van Tulder M, Becker A, Bekkering T, et al. Chapter 3. European guidelines for the management of acute nonspecific low back pain in primary care. Eur Spine J. 2006;15 Suppl 2:S169–191.

32. Page GM, Lacasse A, Quebec Back Pain C, et al. The Quebec Low Back Pain Study: a protocol for an innovative 2-tier provincial cohort. Pain Rep. 2020;5(1):e799.

33. Denis I, Fortin L. Development of a French-Canadian version of the Oswestry Disability Index: cross-cultural adaptation and validation. Spine (Phila Pa 1976). 2012;37(7):E439–444.

34. ThermaCare® Back Pain Therapy. https://www.thermacare.com/heat-wraps/back-pain-therapy. Accessed 2020-07-15.

35. George SZ, Fritz JM, Silfies SP, et al. Interventions for the Management of Acute and Chronic Low Back Pain: Revision 2021. J Orthop Sports Phys Ther. 2021;51(11):CPG1–CPG60.

36. Rolke R, Magerl W, Campbell KA, et al. Quantitative sensory testing: a comprehensive protocol for clinical trials. Eur J Pain. 2006;10(1):77–88.

37. Puta C, Schulz B, Schoeler S, et al. Somatosensory abnormalities for painful and innocuous stimuli at the back and at a site distinct from the region of pain in chronic back pain patients. PLoS One. 2013;8(3):e58885.

38. Arendt-Nielsen L, Yarnitsky D. Experimental and clinical applications of quantitative sensory testing applied to skin, muscles and viscera. J Pain. 2009;10(6):556–572.

39. Georgopoulos V, Akin-Akinyosoye K, Zhang W, McWilliams DF, Hendrick P, Walsh DA. Quantitative sensory testing and predicting outcomes for musculoskeletal pain, disability, and negative affect: a systematic review and meta-analysis. Pain. 2019;160(9):1920–1932.

40. Rolke R, Baron R, Maier C, et al. Quantitative sensory testing in the German Research Network on Neuropathic Pain (DFNS): standardized protocol and reference values. Pain. 2006;123(3):231–243.

41. Bhattacharyya A, Hopkinson LD, Nolet PS, Srbely J. The reliability of pressure pain threshold in individuals with low back or neck pain: a systematic review. Br J Pain. 2023;17(6):579–591.

42. de Oliveira FCL, Cossette C, Mailloux C, Wideman TH, Beaulieu LD, Masse-Alarie H. Within-Session Test-Retest Reliability of Pressure Pain Threshold and Mechanical Temporal Summation in Chronic Low Back Pain. Clin J Pain. 2023;39(5):217–225.

43. Chapman JR, Norvell DC, Hermsmeyer JT, et al. Evaluating common outcomes for measuring treatment success for chronic low back pain. Spine (Phila Pa 1976). 2011;36(21 Suppl):S54–68.

44. Starkweather AR, Heineman A, Storey S, et al. Methods to measure peripheral and central sensitization using quantitative sensory testing: A focus on individuals with low back pain. Appl Nurs Res. 2016;29:237–241.

45. Chen KK, Rolan P, Hutchinson MR, de Zoete RMJ. Reliability of Temporal Summation of Pain in Healthy and Clinical Populations: A Systematic Review and Meta-Analysis. Eur J Pain. 2025;29(8):e70097.

46. Hermens HJ, Freriks B, Disselhorst-Klug C, Rau G. Development of recommendations for SEMG sensors and sensor placement procedures. J Electromyogr Kinesiol. 2000;10(5):361–374.

47. Fraeulin L, Holzgreve F, Brinkbaumer M, et al. Intra- and inter-rater reliability of joint range of motion tests using tape measure, digital inclinometer and inertial motion capturing. PLoS One. 2020;15(12):e0243646.

48. De Carvalho D, Mackey S, To D, et al. A systematic review and meta analysis of measurement properties for the flexion relaxation ratio in people with and without non specific spine pain. Sci Rep. 2024;14(1):3260.

49. Mayer TG, Neblett R, Cohen H, et al. The development and psychometric validation of the central sensitization inventory. Pain Pract. 2012;12(4):276–285.

50. Roelofs J, Peters ML, Patijn J, Schouten EGW, Vlaeyen JWS. Electronic diary assessment of pain-related fear, attention to pain, and pain intensity in chronic low back pain patients. Pain. 2004;112(3):335–342.

51. Wheeler CHB, Williams ACC, Morley SJ. Meta-analysis of the psychometric properties of the Pain Catastrophizing Scale and associations with participant characteristics. Pain. 2019;160(9):1946–1953.

52. Verbeke G, Molenberghs G. Linear Mixed Models for Longitudinal Data. Secaucus, UNITED STATES: Springer; 2000.

53. Nieminen LK, Pyysalo LM, Kankaanpaa MJ. Prognostic factors for pain chronicity in low back pain: a systematic review. Pain Rep. 2021;6(1):e919.

54. Holmberg MJ, Andersen LW. Adjustment for Baseline Characteristics in Randomized Clinical Trials. JAMA. 2022;328(21):2155–2156.

55. Kahan BC, Morris TP. Reporting and analysis of trials using stratified randomisation in leading medical journals: review and reanalysis. BMJ. 2012;345:e5840.

56. Ho F. Regression adjustment for causal inference. BMJ Med. 2025;4(1):e000816.

57. Lluch E, Dueñas L, Falla D, et al. Preoperative Pain Neuroscience Education Combined With Knee Joint Mobilization for Knee Osteoarthritis: A Randomized Controlled Trial. Clin J Pain. 2018;34(1):44–52.

58. Masse-Alarie H, Desgagnes A, Cote-Picard C, et al. Comparisons of the effects of psychologically-informed and usual physiotherapy on pain sensitivity in chronic low back pain: an exploratory randomized controlled trial. Arch Physiother. 2025;15:32–41.

59. Nim CG, Kawchuk GN, Schiøttz-Christensen B, O’Neill S. Changes in pain sensitivity and spinal stiffness in relation to responder status following spinal manipulative therapy in chronic low Back pain: a secondary explorative analysis of a randomized trial. BMC Musculoskeletal Disorders. 2021;22(1):23.

60. Mak JN, Hu Y, Cheng AC, Kwok HY, Chen YH, Luk KD. Flexion-relaxation ratio in sitting: application in low back pain rehabilitation. Spine (Phila Pa 1976). 2010;35(16):1532–1538.

61. Shahvarpour A, Henry SM, Preuss R, Mecheri H, Larivière C. The effect of an 8-week stabilization exercise program on the lumbopelvic rhythm and flexion-relaxation phenomenon. Clin Biomech (Bristol). 2017;48:1–8.

62. Ippersiel P, Cote-Picard C, Roy JS, Masse-Alarie H. Subgrouping People With Acute Low Back Pain Based on Psychological, Sensory, and Motor Characteristics: A Cross-Sectional Study. Eur J Pain. 2025;29(4):e70006.

63. Osumi M, Sumitani M, Otake Y, et al. Kinesiophobia modulates lumbar movements in people with chronic low back pain: a kinematic analysis of lumbar bending and returning movement. Eur Spine J. 2019;28(7):1572–1578.

64. Rose-Dulcina K, Genevay S, Armand S. The flexion-relaxation phenomenon is related to trunk range of motion in low back pain patients: An immersive virtual reality study. Gait & Posture. 2022;97:S225–S226.

