## Supplementary table 1 for "Sensorimotor effects of heatwrap and exercise in acute low back pain: results of a randomised controlled trial"

**Supplementary Table 1.** Group and overall mean and 95% confidence interval of pain sensitivity, flexion-relaxation ratios, current pain intensity, and amplitude of global trunk flexion at each time-points

| Outcomes | **Heatwrap and exercise** | **Heatwrap alone** | **Sham heatwrap** | **Overall** |
| --- | --- | --- | --- | --- |
| Temporal summation of pain (NPRS) | | | | |
| Lumbar | | | | |
| T_0_ | 2.49 (2.02 to 2.96) | 2.07 (1.59 to 2.54) | 2.13 (1.64 to 2.63) | 2.23 (1.95 to 2.50) |
| T_1_ | 3.18 (2.71 to 3.65) | 2.29 (1.82 to 2.77) | 2.69 (2.20 to 3.19) | 2.72 (2.45 to 3.00) |
| T_2_ | 2.78 (2.30 to 3.26) | 2.03 (1.55 to 2.51) | 2.37 (1.87 to 2.87) | 2.39 (2.11 to 2.67) |
| Foot | | | | |
| T_0_ | 2.58 (2.11 to 3.05) | 2.12 (1.64 to 2.60) | 2.10 (1.61 to 2.59) | 2.27 (1.99 to 2.54) |
| T_1_ | 3.13 (2.66 to 3.60) | 2.25 (1.77 to 2.72) | 2.39 (1.90 to 2.88) | 2.59 (2.31 to 2.86) |
| T_2_ | 2.47 (1.99 to 2.95) | 1.66 (1.18 to 2.14) | 2.06 (1.56 to 2.56) | 2.07 (1.78 to 2.35) |
| Pressure pain threshold (kg/cm^2^) | | | | |
| Lumbar | | | | |
| T_0_ | 4.12 (3.34 to 4.90) | 4.34 (3.55 to 5.12) | 4.37 (3.57 to 5.17) | 4.28 (3.82 to 4.73) |
| T_1_ | 4.00 (3.23 to 4.78) | 4.51 (3.72 to 5.30) | 4.53 (3.72 to 5.33) | 4.35 (3.89 to 4.80) |
| T_2_ | 4.38 (3.59 to 5.16) | 5.01 (4.22 to 5.80) | 4.77 (3.97 to 5.58) | 4.72 (4.26 to 5.18) |
| Tibialis anterior | | | | |
| T_0_ | 4.77 (3.97 to 5.58) | 5.01 (4.20 to 5.83) | 4.97 (4.14 to 5.79) | 4.92 (4.45 to 5.39) |
| T_1_ | 4.78 (3.98 to 5.58) | 4.96 (4.15 to 5.77) | 4.68 (3.85 to 5.51) | 4.81 (4.34 to 5.28) |
| T_2_ | 5.28 (4.47 to 6.09) | 5.28 (4.46 to 6.10) | 5.18 (4.35 to 6.01) | 5.25 (4.78 to 5.72) |
| FRR | | | | |
| T12 | | | | |
| T_0_ | 0.53 (0.45 to 0.61) | 0.59 (0.51 to 0.67) | 0.57 (0.49 to 0.66) | 0.56 (0.52 to 0.61) |
| T_1_ | 0.50 (0.42 to 0.58) | 0.57 (0.49 to 0.65) | 0.57 (0.49 to 0.65) | 0.55 (0.50 to 0.59) |
| T_2_ | 0.50 (0.42 to 0.58) | 0.56 (0.48 to 0.64) | 0.59 (0.51 to 0.67) | 0.55 (0.50 to 0.60) |
| L5 | | | | |
| T_0_ | 0.64 (0.55 to 0.73) | 0.60 (0.51 to 0.69) | 0.66 (0.57 to 0.75) | 0.63 (0.58 to 0.69) |
| T_1_ | 0.63 (0.54 to 0.72) | 0.60 (0.51 to 0.69) | 0.67 (0.58 to 0.76) | 0.63 (0.58 to 0.69) |
| T_2_ | 0.58 (0.49 to 0.67) | 0.60 (0.51 to 0.69) | 0.67 (0.57 to 0.76) | 0.62 (0.56 to 0.67) |
| Current pain intensity (NPRS) | | | | |
| T_0_ | 2.86 (2.24 to 3.48) | 2.93 (2.30 to 3.55) | 2.91 (2.27 to 3.55) | 2.90 (2.54 to 3.26) |
| T_1_ | 2.36 (1.74 to 2.98) | 2.31 (1.68 to 2.93) | 2.47 (1.83 to 3.11) | 2.38 (2.02 to 2.74) |
| T_2_ | 1.51 (0.88 to 2.15) | 1.41 (0.77 to 2.05) | 1.15 (0.50 to 1.80) | 1.36 (0.99 to 1.73) |
| ROM – distance from 3^rd^ finger to floor (cm) | | | | |
| T_0_ | 9.99 (6.05 to 13.93) | 12.36 (8.36 to 16.35) | 14.09 (10.03 to 18.15) | 12.15 (9.84 to 14.45) |
| T_1_ | 9.31 (5.36 to 13.26) | 11.96 (7.97 to 15.95) | 12.64 (8.58 to 16.70) | 11.30 (9.00 to 13.61) |
| T_2_ | 6.53 (2.57 to 10.49) | 9.90 (5.89 to 13.89) | 10.57 (6.51 to 14.64) | 9.00 (6.69 to 11.31) |
| T_0_, baseline; T_1_, 1-hour follow-up; T_2_, 1-week follow-up; NPRS, Numerical Pain Rating Scale; FRR, flexion-relaxation ratio; ROM, range of motion (distance from 3^rd^ finger to floor during full trunk flexion). | | | | |
